# Barriers and Facilitators to the Delivery of Age-friendly Health Services in Primary Health Care Centres in Southwest, Nigeria: A Qualitative Study

**DOI:** 10.1101/2023.07.02.23292141

**Authors:** AO Ogunyemi, MR Balogun, AE Ojo, SB Welch, OO Onasanya, VO Yesufu, AT Omotayo, LR Hirschhorn

## Abstract

**Background:** With the rapid growth of Nigeria’s older population, it has become important to establish age-friendly healthcare systems that support care for older people. This study aimed to explore the barriers and facilitators to the delivery of age-friendly health services from the perspectives of primary healthcare managers in Lagos State, Nigeria.

**Method:** We conducted 13 key informant interviews including medical officers of health, principal officers of the PHC Board and board members at the state level. Using a grounded theory approach, qualitative data analysis was initially done by rapid thematic analysis followed by constant comparative analysis using Dedoose software to create a codebook. Three teams of two coders each blind-coded the interviews, resolved coding discrepancies, and reviewed excerpts by code to extract themes.

**Results:** The main barriers to the delivery of age-friendly services included the lack of recognition of older adults as a priority population group; absence of PHC policies targeted to serve older adults specifically; limited training in care of older adults; lack of dedicated funding for care services for older adults and data disaggregated by age to drive decision-making. Key facilitators included an acknowledged mission of the PHCs to provide services for all ages; opportunities for the enhancement of older adult care; availability of a new building template that supports facility design which is more age-friendly; access to basic health care funds; and a positive attitude towards capacity building for existing workforce.

**Conclusion:** While we identified a number of challenges, these offer opportunities to strengthen and prioritize services for older adults in PHCs and build on existing facilitators. Work is needed to identify and test interventions to overcome these challenges and improve the responsiveness of the PHC system to older adults through the delivery of age-friendly health services in PHCs in Lagos, Nigeria.

## Introduction

Age-friendly services are health services that address the unique needs and wants of older people so they can live healthy and active lives [1]. Presently, 14.8 million older persons live in Nigeria and at a growth rate of 3.2%, this number is expected to increase to 30 million by 2050 [2]. Associated with the aging populations, non-communicable diseases (NCDs) have become a leading cause of morbidity, disability and mortality in all regions of the world, including in developing countries [3]. The burden of disease also includes age-related comorbidities including frailty, cognitive decline and multimorbidity [4]. In response, health care services need to be structured to be responsive to the needs of older adults to provide longitudinal primary care to prevent, diagnose and manage these comorbidities and improve the quality of life for this growing population [5, 6].

Primary Health Care centers (PHCs) are well positioned to provide person-centred and community-based care required to prevent or postpone morbidities associated with or more common with aging and reduce their impact on individuals, facilities and health systems [3, 7]. However current PHC-related services in Nigeria prioritize maternal and child healthcare, resulting in gaps in the structure and delivery of services needed by older adults [8, 9]. For example, despite a national action plan for the control of NCDs, its prevention, care and treatment are not currently included in the minimum service package for primary health care in Nigeria [6]. According to the World Health Organization (WHO), PHCs that are age-friendly should address three areas in their care for older adults: 1) information, education, communication and training for providers; 2) health care management systems; and 3) the physical environment [10]. PHCs should focus on adapting health care to the preferences and needs of older people, health promotion and the prevention of chronic conditions that result from aging [11]. These adaptations in service provision can empower older adults, involving them as partners in the care process, with goals to improve their quality of life, and ensure dignity in care [12].

Some studies have identified enablers and barriers to providing age-friendly services in both high and low and middle income countries. For example, in an Australian health system, stakeholders highlighted barriers in delivering hospitals-based education and skills training; governance structures and leadership and working in multidisciplinary teams [13]. Enablers included valuing care of older people and their families; skilled compassionate staff working in effective teams; and care models and environments supporting older people across the system. Focus group data from clinician stakeholders in Canada reflected organizational level barriers and facilitators in five categories namely; education, environment, staffing, policies and other research projects [14]. In Uganda, low readiness for public health facilities to provide age-friendly care services were due to gaps in all of the health system building blocks; leadership, financing, human resources and health management information systems (HMIS) [15].

In Nigeria, high staff workload, administrative challenges, and mismatch with the needs of the aging patient population and existing resources and priorities have been identified as common barriers to service provision for chronic conditions in the PHCs [16]. Minimizing barriers to accessing primary health care and improving care provided for older adults would both enhance health outcomes, and reduce public health spending substantially [12]. Our preliminary data in comprehensive PHCs across Lagos, southwest Nigeria, found health care workers to be inadequately trained for the care of older persons, sub-optimal screening for conditions common among this population, and infrastructural gaps in the physical environment that should allow older people to move around independently, actively, safely and securely while receiving health care (personal communication). To gain further insight into our findings, we used qualitative methods to explore further the barriers and facilitators to the delivery of age-friendly health services from the perspectives of PHC managers. These finding are important to inform the designing and testing of interventions to increase accessibility, acceptability and appropriateness of age-friendly PHCs in Lagos and in the region.

## Materials and methods

### Study design

We conducted a descriptive qualitative study and used the Consolidated criteria for Reporting Qualitative research (COREQ) checklist to report on our methods and finding [17].

### Study site and participants

Lagos State is the most populous city in Africa with over 15 million residents across 57 local districts and 376 wards[18, 19]. There are over 300 PHCs across the wards and 57 of such PHCs are referred to as “flagship PHCs” because they have a full complement of health workers including medical doctors and offer 24-hour services [19]. The Lagos State Primary Health Care Board (PHCB) is tasked with the overall responsibility of primary health care management in the state. The board acts as a strategic link between the state and local councils in matters relating to primary health service improvement and delivery [20]. It is headed by the permanent secretary and who provides oversight functions through the principal officers of several directorates and the medical officers of health (MOH). The MOH oversees all the PHCs within a local council and each PHC facility is headed by an officer-in-charge.

This study was conducted among key informants recruited between May and September 2022. They included medical officers of health, principal officers of the PHC Board who are assistant directors or directors and board members. Participants were purposively selected based on their experience and position in the PHC system. Seventeen individuals who had direct oversight in PHC management and delivery were contacted via invitation letters and phone calls and asked to participate in the study; four declined due to lack of interest and a total of 13 stakeholders were interviewed. However, saturation of themes was achieved in our sample. The demographic characteristics of participants are included in **Table 1**.

**Table 1:**
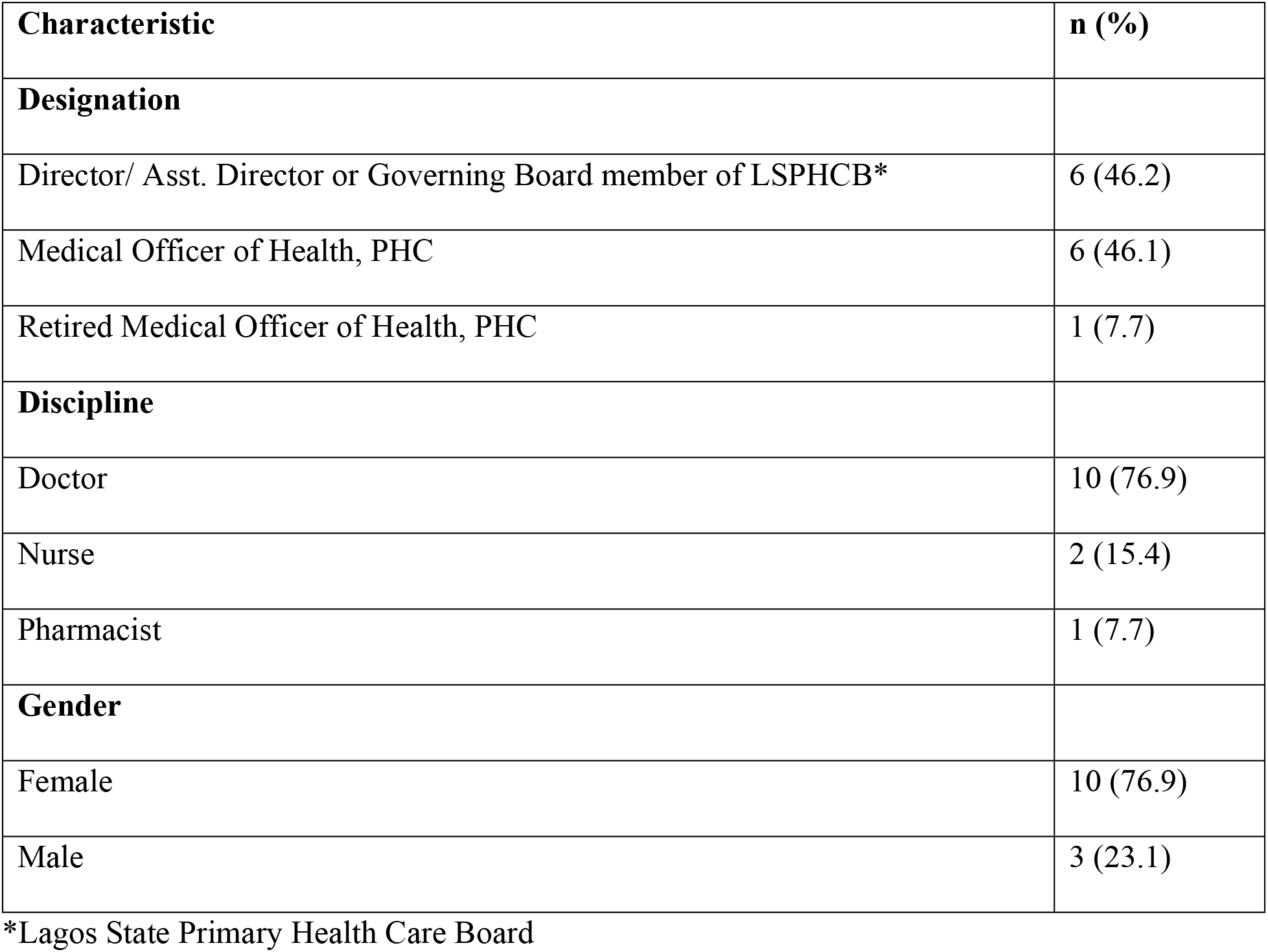
Characteristics of interview participants (n=13)

| Characteristic | n (%) |
| --- | --- |
| **Designation** |  |
| Director/ Asst. Director or Governing Board member of LSPHCB* | 6 (46.2) |
| Medical Officer of Health, PHC | 6 (46.1) |
| Retired Medical Officer of Health, PHC | 1 (7.7) |
| **Discipline** |  |
| Doctor | 10 (76.9) |
| Nurse | 2 (15.4) |
| Pharmacist | 1 (7.7) |
| **Gender** |  |
| Female | 10 (76.9) |
| Male | 3 (23.1) |

\*Lagos State Primary Health Care Board

### Interviewer team

Interviews were conducted by A.O. and M.B., who are both public health specialists with qualitative research experience. Most of the participants were known to the interviewers as a result of the meeting in other public health meetings and related activities in the state. The participants knew the researchers were faculty members at the University of Lagos and were involved in public health research.

### Data collection

The interviews were done virtually at a time convenient to the participants, in a quiet place, using the Zoom platform (Zoom Video Communications, San Jose, CA, USA). At the start of the interview, the interviewer introduced herself, requested permission to record the session and informed verbal consent was obtained as part of the audio recording. Anonymity was ensured by not collecting and including data that could identify the participant during or after the interview. In addition, the participant was informed that a survey on the age-friendliness of services had been undertaken at selected PHCs. The interview began with defining ‘healthy aging’ according to WHO and asking the participants to express their thoughts on what it meant to them. There were nine questions in all, and probing questions were used if required to elicit further information (Suppl. File 1). Interviews lasted about 45 minutes.

All interviews were digitally recorded using the recording feature on the Zoom platform. Once the recording was completed, a separate audio file was used for each participant and saved with a unique identifier in a computer secured by a password known only to the interviewer. Each audio file was transcribed verbatim by a transcription service provider, anonymized and crosschecked by A.O. The transcripts were uploaded to Dedoose (version 9.0.54) by A.O. and made accessible to six coders.

### Data analysis

This study used a grounded theory approach to investigate the research [21]. We used the constant comparative method to analyze our data using an inductive approach.

### Codebook creation

Six coders independently open-coded three transcripts (2 coders per transcript) assigning codes to all concepts identified in the interview data. Through a series of meetings, the coding team condensed the codes to combine similar codes. The resulting codes were discussed and finalized at an all-team meeting with the coders and the PIs. The final codebook (Suppl. File 2) was uploaded into Dedoose.

### Coding

Coders worked in teams of two to code 4-5 transcripts per pair. Teams individually identified and asynchronously discussed discordant code applications through a tracking spreadsheet. Consensus was reached between the pair of coders and the code was changed to reflect the agreed-upon code and placement. If the coding team could not come to agreement, the code discrepancy was discussed by the entire team during a team meeting. Once initial coding was complete all excerpts were reviewed by code, summarized and organized into themes reflecting the barriers and facilitators to implementing age-friendly health services in PHCs. SW led this process through weekly discussions with the coding team.

### Participant Feedback

Preliminary findings as well as emerging findings from a rapid analysis conducted by SW, LH and AO were presented to interviewed participants before the conclusion of analysis. After initial overview, participants were asked if they accurately represented their experience and understanding of the facilitators and barriers to implementing age-friendly health services in PHCs. Feedback from this meeting was used to check the themes developed in the final analysis. There were no additional themes which were identified based on the feedback.

### Ethical Considerations

We obtained ethical approval from the Health Research and Ethics Committee (HREC) of the College of Medicine, University of Lagos (CMUL/HREC/10/21/954). Informed consent was obtained from each participant and confidentiality was ensured through de-identification of transcripts and use of identification codes.

## Results

Three-quarters of the participants were female (76.9%) and doctors (76.9%) (Table 1). Almost half (46.1%) were medical officers of health of the primary health care centers while 46.2% belonged to the management category in the primary health care board.

We identified fifteen major themes (Table 2). Ten of these were barriers to the delivery of age-friendly services while five were facilitators.

**Table 2:**
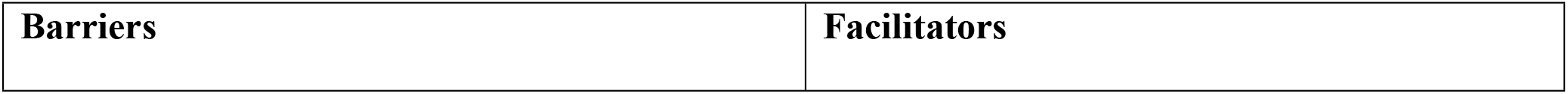
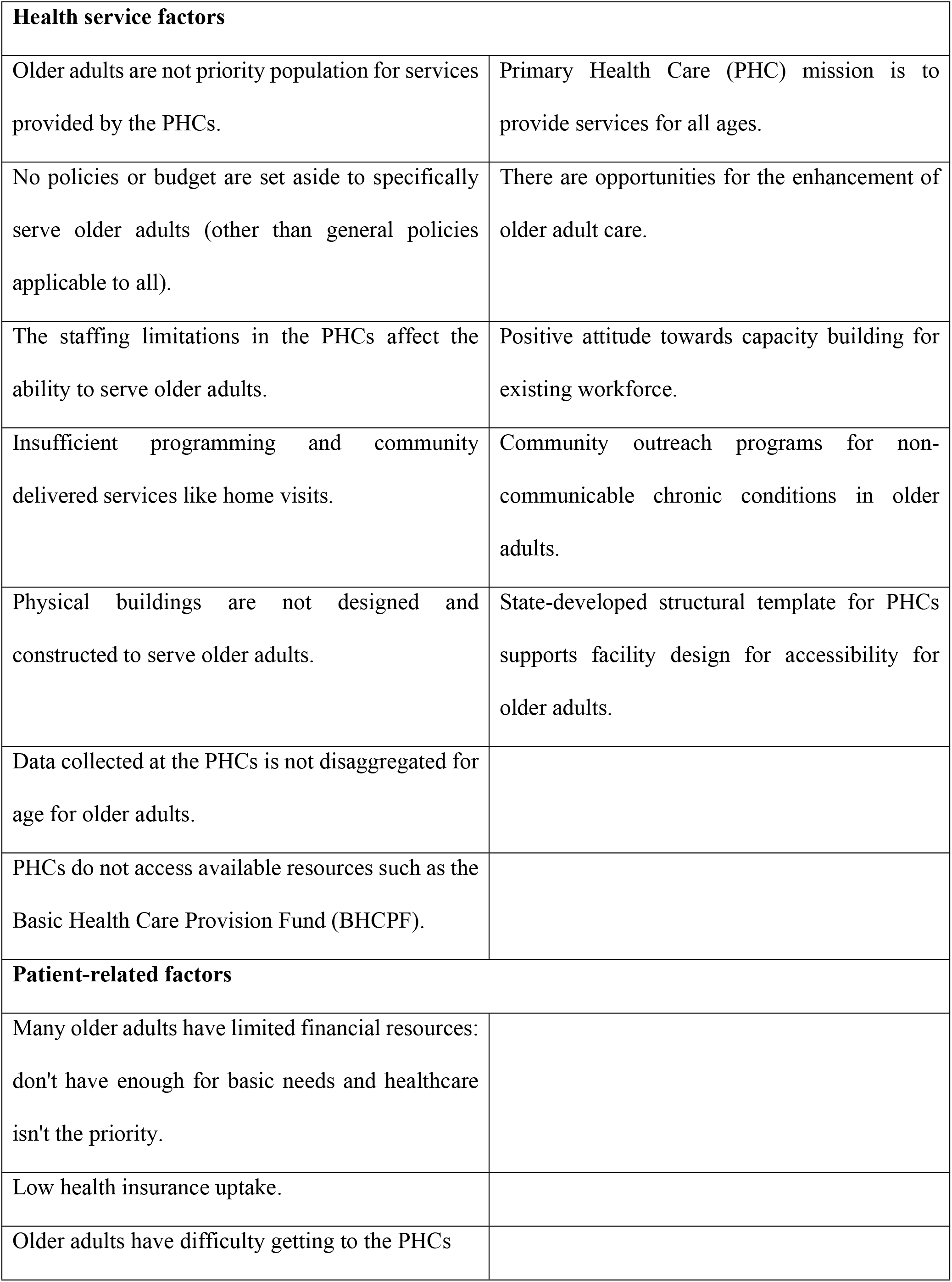
Summary of interview themes: Barriers and facilitators to the delivery of age-friendly health services (AFHS)

| **Health service factors** |  |
| --- | --- |
| Older adults are not priority population for services provided by the PHCs. | Primary Health Care (PHC) mission is to provide services for all ages. |
| No policies or budget are set aside to specifically serve older adults (other than general policies applicable to all). | There are opportunities for the enhancement of older adult care. |
| The staffing limitations in the PHCs affect the ability to serve older adults. | Positive attitude towards capacity building for existing workforce. |
| Insufficient programming and community delivered services like home visits. | Community outreach programs for non-communicable chronic conditions in older adults. |
| Physical buildings are not designed and constructed to serve older adults. | State-developed structural template for PHCs supports facility design for accessibility for older adults. |
| Data collected at the PHCs is not disaggregated for age for older adults. |  |
| PHCs do not access available resources such as the Basic Health Care Provision Fund (BHCPF). |  |
| **Patient-related factors** |  |
| Many older adults have limited financial resources: don't have enough for basic needs and healthcare isn't the priority. |  |
| Low health insurance uptake. |  |
| Older adults have difficulty getting to the PHCs |  |

### Barriers

#### Older adults are not priority population for services provided by the PHCs

There was acknowledgment by the participants that the responsibility of PHC was to provide care across the lifespan and all ages. However, it was noted that older adults were not always a priority population. One of the participants mentioned that Nigeria’s low life expectancy indicates that the focus should be on the younger generation.

> *“Life expectancy is between age 47 to 54 years in Nigeria, so we are more concerned about keeping the younger generation alive at the PHCs. Communicable diseases, treatments of malaria, HIV, TB, those are the bane of our existence in Africa, in Nigeria specifically, so more of the focus is on that.”* P11

> *“We focus on antenatal, we focus on child welfare, we focus on maybe family planning more than the elderly, we focus on those aspects of health than the elderly.” –* P13

One participant noted that older adults should be given preference for receiving services, but it was not always the case.

> *“When elderly people come to the PHCs, we need to give them some priority. We are supposed to give them some priority which we don’t do”* – P13

#### No policies or budget are set aside to specifically serve older adults

Participants expressed that there were no policies that targeted the specific care of older people at the PHC level. In addition, there was no dedicated funding for older adults included in the budget for PHCs.

> *“Unfortunately, I don’t think there’s any policy that is tailored to the aged, of course, there are policies where we talk about equity, and not in particular the aged but that bears in mind all human beings, whether you are young or old, a man or woman, and that’s all encompassing but there’s no policy focusing on the aged, there’s no written policy.”* – P2

> *“To the best of my knowledge there’s no separate financial scheme for the service delivery to older people in the PHCs”*-P6

#### The staffing limitations in the PHCs affect the ability to serve older adults

Staffing limitations was one of the reasons given as a barrier to the provision of dedicated services to older adults at the PHCs. These limitations were said to also impact negatively on providing care to other age groups and various programs, but even more so for older adults given the complexity of care.

> *“Unfortunately, the health worker shortage is a general problem, the fact that we don’t have enough health workers, it cuts across board not only for the elderly but including other age group, other programs…, I really don’t see it feasible currently to have dedicated health workers to the elderly because even what we have is not enough.”* - P2

Knowledge and skill of health workers were considered important by the participants. Another challenge identified was the need for training in the care of older individuals as the health workers available were not trained to manage the needs of older adults resulting in a poor capacity to deliver age-friendly services.

> *“Really, I think my own concern is that we need trained personnel … let’s just do some orientation and reorientation. It’s not easy handling [older] people…you can’t just jump in and do what you think is right, there has to be a structured kind of care or level of care to look after [older people] and minus having doctors occasionally.”* – P3

Participants also stated that there were limitations to services PHCs could offer older adults as some medical services required by them were outside their scope due to staffing expertise.

> *“There is a limit to the services that we can offer. So, it is not a case of not making them a priority. Okay, an elderly that has a very bad diabetes, I am just using that as an instance or uncontrolled blood sugar, there is a limit to what we can do, we don’t have consultants.”* – P12

#### Insufficient programming and community-delivered services

There was a general recognition of the need for community-delivered services such as home visits for older adults. One reason given included varying levels of physical or mental functionality in this age group.

> *“Like I said, for them it will mainly be home visits.. and give them a kind of support, because most times, they are frail and there’s a lack of cognitive functionality, they rarely want to leave home so we need to have scheduled home visits for such older people,”* – P11

Respondents also stressed the importance of community-delivered services to allow for prompt attention to older adults in emergencies and to prevent them from having further complications as well as improving accessibility.

> *“So, these people should have a way of getting care even when it is needed at the community level including emergency… Linking their care right from the community to the PHC and to the secondary level of care should be strengthened so that they can enjoy those services”* – P10

#### Physical buildings are not designed and constructed to serve older adults

Participants discussed challenges in the physical accessibility of the PHCs, for example, the entrances to many of the PHCs did not have ramps as they were not purpose-built.

> *“We can’t say we don’t want this place, you just have to make use of what is being given, the way it’s being built is not accessible or convenient for a frail person to climb the stairs or for a weak person to come in, some are built in places that’s not accessible, bad road structure, no water access. .”*- P2

#### PHCs do not access available resources

The PHCs had not started accessing the Basic Health Care Provision Fund (BHCPF) provided by the federal government which could be helpful in funding the care of the older population.

> *“.. the function of advocacy, reminding the Local Government Health Authorities and the Ward Health Committees that when the funds for Basic Health Care Provision Fund (BHCPF) comes they should prioritize the care of the older population, also there are funds coming in from the health insurance scheme in Lagos State (Ilera eko).”*- P6

#### Data collected at the PHCs are not disaggregated by age for older adults

While there was routine data collection at all the PHCs, there was no disaggregation of the data nor analysis for the purpose of monitoring older adults. As a result, the routine data set does not allow for the forecasting and planning needed to meet the needs of older people. The data collected were mainly used in determining the disease trends and deciding the types of drugs to purchase.

> *“…we can see that people are coming more for this and for that but not in particular having the elderly in mind unfortunately, but generally we look at those data and analyze it to find out if there’s anything different from any trend or anything out of what we are expecting”.* - P2

#### Financial barrier: Many older adults don’t have enough funds even for basic needs, low uptake of insurance

Participants shared that older adults who had limited funds will have difficulty prioritizing their health or visiting the clinics. This is because certain payments had to be made to access care as well as the need to pay for food, habitation and so on. Some of this challenge is because of the low uptake of the Lagos State health insurance scheme.

> *“Of course, there’s health insurance established in Lagos State but just few people are enrolled on the program, they still have to pay from pocket to get their healthcare and most of them because they are retired do not have funds to get the best services, sometimes they have limited funds and just prioritize the kind of drugs they can buy and these are the main challenges.”*- P2

> *“The challenges will be, people are not taking up even the health insurance mandatory program, people are not taking, they are not buying the insurance”* - P12

#### Older adults have difficulty getting to the PHCs

The participants noted that there were challenges in accessing the PHCs including the needed social support and transport to bring older adults to the PHCs.

> *“The only challenge I feel we have is if they don’t have people around them to support them to the health facility”*- P5

Geographic barriers including topography and distance to existing PHCs were among other reasons why older adults were not accessing some of the PHCs.

> *“…some chairmen will just build PHCs even at places that are very difficult to reach and eventually nobody is ready to go maybe because of the topography of the area, you know the hill and the rest and some are not because of the road structure, some are very difficult to access because of these things we are talking about.”* – P4

### Facilitators

We heard a number of facilitators from respondents on the capacity to provide age friendly care at the PHC.

#### Primary Health Care mission is to provide services for all ages

Many of the participants acknowledged that it was the role of the PHCs to provide affordable care across the lifespan and therefore it is open to all including older adults.

> *“At the primary health care centre,…. we provide services that might not be targeted at ageing specifically but it’s a general service…. So even though it’s not targeted, it is open to people of all ages.”* -P11

#### There are opportunities for the strengthening care for older adult

The participants mentioned that there were opportunities within the PHC system to strengthen care for older adults. The Basic Health Care Provision Fund (BHCPF) by the federal government and the Lagos State Health Scheme (LSHS) are available to the PHCs, although as noted above, there was inadequate uptake by PHCs.

> *“If more PHCs are set and enrolled in the program and we have people coming into the insurance program, then it will be easy, because this is a very good opportunity in the state and there is a lot of money going into it, there’s so much funding coming from the basic healthcare provision funds…. ”* - P2

Availability of Social and welfare services in some areas such as nutritional support, money, and prescriptions were also identified as important to help address the healthcare needs of older adults.

> *“There are services that are good for elderly people but are not directed towards health, there are some local governments, who have nutrition services for elderly people, and some give financial transfers to support them”*- P1

#### Positive attitude towards capacity building for existing workforce

Some of the participants felt that capacity building of the existing workforce to expand and enhance the provision of age-friendly services could be done through training. An example given was the involvement of community mobilizers to encourage older adults who needed th care to access the PHCs and also in the provision of some home services.

> *“We also have social mobilizers and these are people that have been recognized as people that have the voice within the community, they have been trained over the years, they go round talk to people and they’ll share materials available like posters, fliers, handbills. Where and how to behave in the community and where they can ask questions, sometimes it’s mostly in form of meetings”* - P2

#### Community outreach programs for non-communicable chronic conditions in older adults

PHCs already provide screening and treatment during community outreach programs for common chronic conditions such as hypertension and diabetes. Although they do not routinely manage these conditions in the PHCs, it can be built upon over time.

> *“We’ve actually been doing several care for elderly, on mostly non communicable diseases like hypertension and diabetes, there are times when we have scheduled [community] programs like hypertension week, wellness week”*- P2

#### State-developed structural template for PHCs supports facility design for accessibility for older adults

The participants reported that Lagos State has a building template which reflects many of the WHO Age-Friendly guidelines and that all new PHC construction must follow this template. They also noted that it was possible for PHCs to use their quarterly funds to make the changes needed to their buildings. They expressed that with the political will, funds were available and the PHCs just needed to know what to do.

> *“It’s not hard. So far there is a political will, you know political will, it will bring out the funds, the Medical Officer of Health (MOH) can only make recommendation that are okay for the elderly…., you need all these chairs with armrest, high chairs not the low ones with backrest and armrest and then we need a ramp both in front and at the back, the ramps for Ibeju is at the side which is meant to be the emergency entrance.”* P9

## Discussion

Our qualitative work found supporting evidence of the earlier quantitative findings of gaps in training and infrastructure, as well as identified new barriers to providing healthy aging care for older people in Lagos state as well as the presence of some facilitators. Our respondents highlighted that older adults were not always a priority population for care at the PHCs but acknowledged the centrality of PHC’s role to provide accessible and affordable care across the lifespan. While the need to strengthen accessibility as well as the availability of PHC focused on older adults was recognized there were a number of challenges that need to be addressed.

One important gap noted by some of the respondents was the lack of the existence of policies targeted at serving older adults in PHCs in Lagos State. However, it was stated that there were aspects of the national policies that implied the inclusion of older adults for care based on care for all adults with the main goal of social welfare. For example, according to the 1999 Nigerian Constitution, Section 16, sub-section 2(d), old age care and pensions, as well as sick benefits, will be provided to all citizens [22]. Similarly, existing policy documents in Uganda focus only on addressing the social needs of older adults[15]. The federal government of Nigeria introduced new policies such as the National Senior Citizens Centre in 2017, an agency to cater to the needs of senior citizens; and related matters were enacted and in due course, the National Policy on Ageing for Older Persons in Nigeria was approved by the Federal Executive Council in February 2021[23, 24]. In addition to this, financial barriers from the PHC and limited financial resources among older adults were also identified. These barriers included low rates of uptake of programs designed to reduce these barriers. For example, the respondents also identified the poor coverage by the National Health Insurance Scheme (NHIS), reflections supported by data that only 5% of the population, mostly civil servants and younger individuals were enrolled in the program [25].

Another important barriers was the staffing limitations and specifically workforce trained in the care for older adults was another reason given as an implementation barrier for AFHS. These results reflect similar to findings from a quantitative survey in Benin City, Nigeria [26]. The lack of health manpower capacity is an issue of global public health concern and a major limitation to implementing important health policies across the lifespan [27]. In Nigeria, similar to in many countries in Africa, training in the care of older persons is still new. In Nigeria, it is not included in the undergraduate curriculum and only a few hospitals are accredited to offer training in geriatrics [28]. As a result, healthcare workers often do not have the knowledge and competency to address the emerging challenges in delivering care for older adults living with complex needs and this threatens the quality of care as well as trust in the healthcare system [29].

Many participants also noted the importance of moving beyond facility-based care to providing home services in the community to support older adults but there are no such formalized services for this age group in Nigeria [30]. This work would be important for both practice outreach as well as prompt attention to identifying and responding to emergency situations, as well as active outreach to prevent delay in disease complications. Learning from existing approaches that work will be important as well as leveraging strengths in community care. For example, in parts of the United States, community-based supports and services are designed to help community-dwelling older adults remain safely in their homes and delay the need for placing them in institutions [31]. Respondents also noted that many older adults lacked the social support in bringing older adults to the facility was also identified as a challenge. Studies have suggested that some of this weakened social support and this has been largely attributed to the change in family dynamics and traditional family structure in Nigeria as well as the lack of community-based support noted above [32].

An important barrier reported was that many of the PHCs were not designed and constructed to serve older adults, especially as regards physical accessibility. For example, some of the buildings that serve as PHCs were put up by the local council chairmen for the purpose of providing care in the community, and health providers regarded this as a starting point. In our preliminary study that assessed the facility readiness of selected PHCs for AFHS, we found that two-fifths of the PHCs did not have ramps at their entrances, and a third did not have a reception area near the entrance. In addition, 86.7% of the public toilets in the PHCs did not have grab bars, and 33.3% were not served by public transportation. According to several of the participants, there was a current effort by the present state government to ensure donors complied with the building template provided. Furthermore, the participants were optimistic that funds were not a barrier to making recommended changes to older buildings if given the necessary information and expanded use of the available funding sources including increasing enrollment in insurance and the BPHFC funds. Among the three principles proposed by WHO for AFHS, the physical environment has the most precise guidelines and is thought to be the least complex area of change [11].

While data are routinely recorded and reported at the PHCs, it was reported that they were not disaggregated for age for the older adult category, 60 years and above. One of our participants mentioned that the nationally designed reporting forms did not specify for those aged 60 years and above but classified adults as 18 years and above. This negatively affected the use of data to plan for the needs of older adults in the PHCs. However, routinely collected data is used to understand disease trends in the community and to restock drugs for facility use. On the contrary, outpatient department registers in Uganda had disaggregated data by age in all the selected health facilities in a study but geriatric data was not being used to improve service delivery [15]. A multi-country study on health data reporting in PHCs in five LMICs including Nigeria found that data was primarily collected in outpatient care, antenatal care, immunization, family planning, HIV and tuberculosis services only [33]. Although older adults vary greatly in terms of health status, the majority of them have at least one chronic condition and are prone to other age-related diseases such that the current data reporting system in the PHCs will exclude them from care [34, 35]. In comparison to high-income countries, data on aging and the well-being of older people in LMICs are insufficient and might not be fully representative of the aging populations in the countries flagging the need to prioritize research for informed decision-making [36].

One of the key facilitators for the implementation of AFHS mentioned by our respondents was the establishment of the Basic Healthcare Provision Fund (BHCPF), a component of the National Health Act of 2014. It is predominantly funded by the Federal Government of not less than 1% of the Consolidated Revenue Fund (CRF), donor grants, and others [37]. Our respondents were positive that accessing the BHCPF would provide opportunities for investments in the PHCs but admitted that such funding may still not be targeted toward the care of older people. However this was also identified as missed opportunity identified was low use of the Basic Healthcare Provision Fund (BHCPF), PHCs which could support expanding and strengthening care for older adults. Our findings are supported by a study of the process of administration and disbursement of the BHCPF which highlighted some challenges such as poor accountability systems, high-level political interference, and lack of political commitment from the State to release funds for health activities[38].

Another facilitator mentioned by our participants while reflecting on the success of other PHC programs, is the existing PHC workforce that can be trained on the delivery of AFHS. This includes community health workers who can provide health awareness campaigns and home services for older adults while performing their routine duties in the community. Community health workers constitute the majority of the health workers in PHCs and their deployment to deliver essential health services has proven to be a well-established strategy to address the critical skilled health workforce [39, 40]. Several respondents also mentioned existing community outreach programs for non-communicable chronic conditions in older adults by the PHCs as a facilitator. Such care is offered during occasional outreaches comprising free screening and treatment of chronic conditions such as hypertension and diabetes. Recently, the Nigerian government focused on PHCs to tackle the rising non-communicable disease (NCD) burden by strengthening community health workers’ skills and competencies for NCD care delivery [40]. However, it has been noted that such roles are sometimes beyond the community health worker’s scope of practice and may be limited by access to adequate training and supervision [40].

To the best of our knowledge, this study is the first to explore barriers and facilitators to the delivery of age-friendly health services in Nigeria and one of few from Africa. We interviewed a multidisciplinary group of participants at high level of leadership in the PHC setting. Our studies had a number of limitations. Like all qualitative methods, results reflect the opinion and reflections of the respondents and their possible biases. We worked to limit bias by keeping questions open ended and ensuring anonymity throughout the process. Importantly we did not include older adults or their care givers in this study due to resource limitations, so their important perspectives and experiences of care were not captured. However we are planning to engage these groups in a follow-on study

## Conclusions

In conclusion, this study among PHC stakeholders recognizes the role of PHCs in the promotion of healthy aging in general. While barriers were identified.. there were multiple many opportunities to strengthen and prioritize primary care services for older adults in Lagos state. Actions are needed to further engage stakeholders to prioritize actions and co-design and implement priority interventions that will directly address the barriers and build on exisiting strengths to improve the delivery of age-friendly health services in PHCs in Lagos, Nigeria.

## Data Availability

Data cannot be shared publicly because of the possibility of violating participantș;s privacy. Data are available from the Secretary, Health Research Ethics Committee, Institutional Data Access / Ethics Committee (contact via) for researchers who meet the criteria for access to confidential data.

## Acknowledgements

The research reported in this publication was generously supported by the Robert J. Havey, MD Institute for Global Health’s catalyzer fund at Northwestern University, Feinberg School of Medicine. The authors appreciate the funder for the catalyzer grant to conduct this research. The funder had no input in the study findings or interpretation. We also thank all our interview participants for sharing their experiences, granting access to the research team, and for their time in responding to the survey.

## Supporting Information

S1: Qualitative interview guide

S2: Coding tree

S3: Table of additional quotes

## Notes

### Competing Interest Statement

The authors have declared that no competing interests exist.

### Funding Statement

-LRH -(No award number) -Robert J. Havey, MD Institute for Global Healthș;s catalyzer fund at Northwestern University, Feinberg School of Medicine https://www.globalhealth.northwestern.edu/-The funders had no role in study design, data collection and analysis, decision to publish, or preparation of the manuscript.

### Author Declarations

Health Research Ethics Committee, CMUL College of Medicine, University of Lagos Research Management Office Chairperson Prof. S.A. Omilabu; Organisation administrator's name or contact person Hodefe Olufemi; Physical address REC 11 address Postal address CMUL, Health Research Ethics Committee (HREC), c/o Office of the Provost College of Medicine, University of Lagos, Idi-Araba, Lagos, Nigeria. Country Nigeria Institution website address http://cmul.unilag.edu.ng/

## References

1. John A. Hartford Foundation. What is Age-Friendly Care? 2023. Available from: https://www.johnahartford.org/grants-strategy/current-strategies/age-friendly/age-friendly-care.

2. National Senior Citizens Centre. National Plan of Action on Ageing in Nigeria and Project Activities 2021-2025. Abuja, Nigeria2022.

3. World Health Organization. Active Ageing: A Policy Framework 2002 [cited 2023 2nd June]. Available from: https://extranet.who.int/agefriendlyworld/wp-content/uploads/2014/06/WHO-Active-Ageing-Framework.pdf.

4. Kadambi S AM, Loh KP Multimorbidity, Function, and Cognition in Aging. Clin Geriatr Med. 2020;36(4):569-584. doi: 10.1016/j.cger.2020.06.002. PubMed Central PMCID: PMCPMC8012012.

5. World Health Organization. Noncommunicable diseases 2022 [cited 2022 16 September 2022]. Available from: https://www.who.int/news-room/fact-sheets/detail/noncommunicable-diseases.

6. Federal Ministry of Health N. National Multi-Sectoral Action Plan for the Prevention and Control of Non-communicable Diseases (2019–2025) Abuja, Nigeria2019 [cited 2023 3rd February]. Available from: https://www.health.gov.ng/doc/NCDs_Multisectoral_Action_Plan.pdf.

7. Ajisegiri WS, Abimbola S, Tesema AG, Odusanya OO, Peiris D, Joshi R. The organisation of primary health care service delivery for non-communicable diseases in Nigeria: A case-study analysis. PLOS Global Public Health. 2022;2(7):e0000566. doi: 10.1371/journal.pgph.0000566.

8. Okonofua F, Ntoimo LF, Yaya S, Igboin B, Solanke O, Ekwo C, et al. Effect of a multifaceted intervention on the utilisation of primary health for maternal and child health care in rural Nigeria: a quasi-experimental study. BMJ Open. 2022;12(2):e049499. doi: 10.1136/bmjopen-2021-049499.

9. Obembe TA, Osungbade KO, Ibrahim C. Appraisal of primary health care services in Federal Capital Territory, Abuja, Nigeria: how committed are the health workers? Pan Afr Med J. 2017;28:134. Epub 20171011. doi: 10.11604/pamj.2017.28.134.12444. PubMed PMID: 29541284; PubMed Central PMCID: PMCPMC5847130.

10. World Health Organization. Age-friendly Primary Health Care Centres Toolkit [Internet]. WHO. 2008. Available from: https://extranet.who.int/agefriendlyworld/wp-content/uploads/2014/06/WHO-Age-friendly-Primary-Health-Care-toolkit.pdf.

11. Tavares J, Santinha G, Rocha NP. Age-Friendly Health Care: A Systematic Review. Healthcare (Basel). 2021;9(1). Epub 20210116. doi: 10.3390/healthcare9010083. PubMed PMID: 33561084; PubMed Central PMCID: PMCPMC7830866.

12. Gomes ID, Sobreira LS, Da Silva LV, De Souza MCMR, Karina G, Souto RQ. Age-Friendly Primary Health Care: A Scoping Review. Springer International Publishing; 2022. p. 94–107.

13. Mudge AM, Young A, McRae P, Graham F, Whiting E, Hubbard RE. Qualitative analysis of challenges and enablers to providing age friendly hospital care in an Australian health system. BMC Geriatr. 2021;21(1):147. Epub 20210227. doi: 10.1186/s12877-021-02098-w. PubMed PMID: 33639854; PubMed Central PMCID: PMCPMC7913259.

14. Hanson HM, Warkentin L, Wilson R, Sandhu N, Slaughter SE, Khadaroo RG. Facilitators and barriers of change toward an elder-friendly surgical environment: perspectives of clinician stakeholder groups. BMC Health Services Research. 2017;17(1). doi: 10.1186/s12913-017-2481-z.

15. Ssensamba JT, Mukuru M, Nakafeero M, Ssenyonga R, Kiwanuka SN. Health systems readiness to provide geriatric friendly care services in Uganda: a cross-sectional study. BMC Geriatrics. 2019;19(1). doi: 10.1186/s12877-019-1272-2.

16. Odusola AO, Stronks K, Hendriks ME, Schultsz C, Akande T, Osibogun A, et al. Enablers and barriers for implementing high-quality hypertension care in a rural primary care setting in Nigeria: perspectives of primary care staff and health insurance managers. Global Health Action. 2016;9(1):29041. doi: 10.3402/gha.v9.29041.

17. Tong A SP, Craig J. . Consolidated criteria for reporting qualitative research (COREQ): a 32-item checklist for interviews and focus groups. Int J Qual Health Care. 2007;19(6):349– 57.

18. Lagos State Government WebsiteLagos State Government [cited 2022 December 31st]. Available from: http://www.lagosstate.gov.ng/pagelinks.php?p=7.

19. Abosede OA SO. Strengthening the Foundation for Sustainable Primary Health Care Services in Nigeria. . Primary Health Care 2014;4(3). doi: doi:10.4172/2167-1079.1000167.

20. Lagos State Primary Health Care Board. About PHCB [cited 2023 19th June]. Available from: https://lsphcb.lg.gov.ng/about-phcb/.

21. Chun Tie Y, Birks M, Francis K. Grounded theory research: A design framework for novice researchers. SAGE Open Medicine. 2019;7:205031211882292. doi: 10.1177/2050312118822927.

22. Federal Republic of Nigeria. 1999 Nigerian Constitution 1999 [cited 2023 3rd April]. Available from: https://nigerian-constitution.com/.

23. Assembly N. National Senior Citizens Centre Act, 2017 2017 [cited 2023 3rd April]. Available from: https://placng.org/i/wp-content/uploads/2019/12/National-Senior-Citizens-Centre-Act-2017.pdf.

24. National Senior Citizens Centre. NSCC Strategic Roadmap on Ageing. Abuja, Nigeria: National Senior Citizens Centre Head Office; 2022 October 2022. 80 p.

25. Federal Ministry of Health Nigeria. Second national strategic health development plan 2018–2022. Federal Government of Nigeria. 2018 [cited 2023 3rd April]. Available from: http://ngfrepository.org.ng:8080/jspui/handle/123456789/3283.

26. Tanyi PL, Pelser A. Towards an “age-friendly-hospital”: Older persons’ perceptions of an age-friendly hospital environment in Nigeria. Cogent Medicine. 2020;7(1):1853895. doi: 10.1080/2331205x.2020.1853895.

27. Aluko JO, Anthea R, Marie Modeste RR. Manpower capacity and reasons for staff shortage in primary health care maternity centres in Nigeria: a mixed-methods study. BMC Health Services Research. 2019;19(1). doi: 10.1186/s12913-018-3819-x.

28. Yvonne Buowari D. Geriatric Care in Africa. IntechOpen; 2022.

29. Demiris G, Hodgson NA, Sefcik JS, Travers JL, McPhillips MV, Naylor MD. High-value care for older adults with complex care needs: Leveraging nurses as innovators. Nursing Outlook. 2020;68(1):26–32. doi: 10.1016/j.outlook.2019.06.019.

30. Okoye UO. Community-Based Care For Home Bound Elderly Persons In Nigeria; A Policy Option International Journal of Innovative Research in Science, Engineering and Technology 2013;Vol. 2(Issue 12).

31. Siegler EL, Lama SD, Knight MG, Laureano E, Reid MC. Community-Based Supports and Services for Older Adults: A Primer for Clinicians. Journal of Geriatrics. 2015;2015:1–6. doi: 10.1155/2015/678625.

32. Animasahun VJ, Chapman HJ. Psychosocial health challenges of the elderly in Nigeria: a narrative review. African Health Sciences. 2017;17(2):575. doi: 10.4314/ahs.v17i2.35.

33. Siyam A, Ir P, York D, Antwi J, Amponsah F, Rambique O, et al. The burden of recording and reporting health data in primary health care facilities in five low- and lower-middle income countries. BMC Health Services Research. 2021;21(S1). doi: 10.1186/s12913-021-06652-5.

34. Odland ML, Payne C, Witham MD, Siedner MJ, Bärnighausen T, Bountogo M, et al. Epidemiology of multimorbidity in conditions of extreme poverty: a population-based study of older adults in rural Burkina Faso. BMJ Global Health. 2020;5(3):e002096. doi: 10.1136/bmjgh-2019-002096.

35. Faronbi JO, Ademuyiwa IY, Olaogun AA. Patterns of chronic illness among older patients attending a university hospital in Nigeria. Ghana Medical Journal. 2020;54(1):42–7. doi: 10.4314/gmj.v54i1.7.

36. Goodman-Palmer D, Ferriolli E, Gordon AL, Greig C, Hirschhorn LR, Ogunyemi AO, et al. Health and wellbeing of older people in LMICs: a call for research-informed decision making. Lancet Glob Health. 2023;11(2):e191–e2. doi: 10.1016/s2214-109x(22)00546-0. PubMed PMID: 36669801.

37. Federal Ministry of Health Nigeria. Guidelines for the administration, disbursement and monitoring of the basic healthcare provision fund 2020 [cited 2023 3rd April]. Available from: https://www.health.gov.ng/doc/BHCPF-2020-Guidelines.pdf.

38. Uzochukwu B, Onwujekwe E, Mbachu C, Okeke C, Molyneux S, Gilson L. Accountability mechanisms for implementing a health financing option: the case of the basic health care provision fund (BHCPF) in Nigeria. International Journal for Equity in Health. 2018;17(1). doi: 10.1186/s12939-018-0807-z.

39. Olaniran A, Smith H, Unkels R, Bar-Zeev S, Van Den Broek N. Who is a community health worker? – a systematic review of definitions. Global Health Action. 2017;10(1):1272223. doi: 10.1080/16549716.2017.1272223.

40. Ajisegiri WS AS, Tesema AG, Odusanya OO, Peiris D, Joshi R. “We just have to help”: Community health workers’ informal task-shifting and task-sharing practices for hypertension and diabetes care in Nigeria. Front Public Health. 2023;11:1038062. doi: doi:10.3389/fpubh.2023.1038062.

